# Rethinking the pathogenicity of intragenic *DMD* duplications detected by carrier screening: high prevalence of non-tandem duplications revealed by long-read sequencing

**DOI:** 10.1101/2025.04.10.25325596

**Authors:** Qiliang Ding, Jagadheshwar Balan, Noemi Vidal-Folch, Angela M. Pickart, Guangchao Sun, Jesse R. Walsh, Ramanath Majumdar, Eric W. Klee, Stephen J. Murphy, Devin Oglesbee, Ross A. Rowsey, Linda Hasadsri

## Abstract

**Purpose:** The pathogenicity of intragenic duplications depends on their structural configuration. Tandem duplications often disrupt reading frames and cause gene loss-of-function, whereas interspersed (non-tandem) duplications are largely benign. When the configuration cannot be determined, current guidelines presume a tandem structure, leading to some laboratories automatically classifying such variants as likely pathogenic or pathogenic. This study evaluates the validity of this presumption for *DMD*, in patients with and without clinical indications of dystrophinopathy.

**Methods:** We performed high-coverage whole-genome long-read sequencing on 15 patients with intragenic *DMD* duplications. Four patients had clinically indicated dystrophinopathy testing, while in the remaining 11 patients, the duplications were detected without clear indications of dystrophinopathy (e.g., “incidentally detected” through carrier screening).

**Results:** All four patients with clinical indications had tandem duplications. In contrast, 64% (7/11) of the incidentally detected cases had interspersed duplications, with four subsequently re-classified as likely benign, two likely pathogenic, and one uncertain. These duplications were often complex, involving co-duplications or co-deletions with other regions.

**Conclusion:** Our findings challenge the presumption that intragenic *DMD* duplications are predominantly tandem. This highlights the need for a cautious variant interpretation approach, particularly in carrier screening and other settings where variants are identified without indications of dystrophinopathy.

## Introduction

The *DMD* gene is associated with X-linked dystrophinopathies (also referred to as *DMD*-related disorders), including Duchenne muscular dystrophy (DMD), Becker muscular dystrophy (BMD), and *DMD*-associated dilated cardiomyopathy. Patients with DMD who are assigned male at birth (AMAB) typically exhibit muscle weakness in early childhood, loss of ambulation by age 10–12, and death by age 30. Compared with DMD, BMD is a milder disease characterized by later onset, slower progression, and longer life expectancy.^1^

DMD and BMD are relatively common genetic conditions, with a prevalence of 1.38 per 10,000 U.S. AMABs aged 5–24 years.^2^ An Israeli study estimated the carrier frequency of dystrophinopathy in individuals assigned female at birth (AFAB) to be 1 in 1,374.^3^ Due to the carrier frequency and significant disease severity, the American College of Medical Genetics and Genomics (ACMG) recommends offering *DMD* carrier screening to all patients who are pregnant or considering pregnancy.^4^

Intragenic duplications—duplications involving one or more exons but fully contained within a given gene— are a significant source of *DMD* pathogenic variants, with studies attributing 5–20% of dystrophinopathy cases to these variants.^1,5^ Nonetheless, not all intragenic *DMD* duplications are pathogenic. Intragenic duplications can be either tandem or interspersed (also known as non-tandem or insertional). In tandem duplications, the duplicated segment is joined adjacent to the original segment in the same orientation, therefore may disrupt the reading frame and lead to pathogenicity. In contrast, interspersed duplications generally do not disrupt the reading frame and are possibly benign.

The disease-causing intragenic *DMD* duplications found in dystrophinopathy patients are in tandem.^6^ However, the structural configuration (i.e., tandem vs. interspersed) of such variants is not routinely assessed in clinical laboratory practice, partly due to technical limitations. Most current clinical assays for copy-number detection are unable to distinguish between tandem and interspersed duplications. These assays include multiplex ligation-dependent probe amplification (MLPA), chromosomal microarray analysis (CMA), and most next-generation sequencing (NGS) assays. Instead, current variant interpretation guidelines—namely, the application of PVS1^7^ and PM4^8^—presume intragenic duplications are in tandem unless proven otherwise (hereafter referred to as the “tandem presumption”).

While the tandem presumption is likely accurate in patients with a strong clinical indication for dystrophinopathy testing (i.e., personal or family history), little is known about intragenic *DMD* duplications detected in individuals without such an indication (hereafter referred to as “incidentally detected”). This represents a critical knowledge gap, as many patients undergo *DMD* genetic testing as part of carrier screening in the absence of a clinical indication of dystrophinopathy. If the tandem presumption is inaccurate in these patients, it could lead to overestimation of variant pathogenicity—potentially resulting in unnecessary invasive procedures for diagnostic fetal testing and/or even pregnancy termination.

In this study, we performed Oxford Nanopore Technologies (ONT)-based long-read sequencing on samples from 15 patients with intragenic *DMD* duplications. The duplications were in tandem in all four cases with a clinical indication of dystrophinopathy. However, among the 11 incidentally detected cases, seven (64%) involved interspersed duplications. We also developed DMDuper, a computational tool that accurately distinguishes tandem from interspersed intragenic *DMD* duplications using long-read sequencing data, with 100% accuracy across the 15 cases. Taken together, our findings challenge the tandem presumption for incidentally detected intragenic *DMD* duplications (e.g., those identified by carrier screening) and highlight the need for a cautious approach to variant classification in these cases.

## Materials and Methods

DNA was extracted using Qiagen (Hilden, Germany) or Chemagic (Revvity, Waltham, MA) kits, which was then processed with the Short Read Eliminator kit (PacBio, Menlo Park, CA), and subsequently sheared to 30 kb on Megaruptor 3 (Seraing, Belgium). Library was prepared using the SQK-LSK114 kit and sequenced using R10.4.1 flow cells on a PromethION 24 (ONT, Oxford, UK) for 72 h, with one sample per flow cell. Base calling was performed by Dorado version 0.8.0 (https://github.com/nanoporetech/dorado). Alignment to GRCh38 was performed using Winnowmap2 version 2.03 (https://github.com/marbl/Winnowmap).^9^ The Integrative Genomics Viewer (IGV) was used for manual review of the BAM files. Long-range phasing was manually performed using IGV by identifying reads that share the same single-nucleotide variants. The UCSC BLAT tool was used to investigate read clips (https://genome.ucsc.edu/cgi-bin/hgBlat). All genomic coordinates in this manuscript are in GRCh38/hg38.

In addition, we developed DMDuper, a bioinformatics tool that can automatically distinguish between tandem and interspersed duplications. Briefly, DMDuper first recognizes a *DMD* duplication by detecting soft-clipped read signatures at the breakpoints. DMDuper then extracts clipped reads at the breakpoints and performs haplotype-aware assembly using Flye version 2.9.4 (https://github.com/mikolmogorov/Flye). The assembled contigs are mapped to the reference genome using minimap2 version 2.18^10^ (https://github.com/lh3/minimap2), and the alignments are used to infer the configuration. DMDuper is available at https://github.com/jagadhesh89/dmduper.

## Results

We analyzed 15 samples with intragenic *DMD* duplications using ONT high-coverage whole-genome long-read sequencing (see Table S1 for sequencing metrics). Four of the 15 cases had a personal or family history consistent with dystrophinopathy (DMD-1 to DMD-4, Table 1), while the remaining 11 did not have a clear indication of dystrophinopathy (DMD-5 to DMD-15, Table 1). These 11 cases included five ascertained through carrier screening and six that underwent microarray testing for other indications (e.g., congenital heart disease, short stature, autism).

**Table 1.** Clinical and laboratory information of the cases included in this study.

| Case ID | Sex | Reason for testing | Clinical assay and results<sup>1</sup> | Results from long-read sequencing |  |  |
| --- | --- | --- | --- | --- | --- | --- |
|  |  |  |  | Configuration | Breakpoints | Classification |
| DMD-1 | M | Diagnosis of dystrophinopathy | MLPA, exon 2 duplication | Tandem | X:32882909-33047562 | Pathogenic |
| DMD-2 | F | Family history of dystrophinopathy | MLPA, exons 13–17 duplication | Tandem | X:32524934-32613200 | Pathogenic |
| DMD-3 | F | Family history of dystrophinopathy | MLPA, exons 2–7 duplication | Tandem | X:32753880-33032291 | Pathogenic |
| DMD-4 | F | Consistently elevated creatine kinase level | MLPA, exons 6–7 duplication | Tandem | X:32805559-32820064 | Pathogenic |
| DMD-5 | F | Follow-up positive carrier screening | MLPA, exons 45–55 duplication | Tandem | X:31551483-32081542 | Pathogenic |
| DMD-6 | F | Autism | CytoScan HD microarray, arr[hg38] Xp21.1(32,740,659-32,814,457)x3 | Tandem | X:32743240-32814624 | Pathogenic |
| DMD-7 | F | Carrier screening | GeneTitan genotyping microarray, exon 42 duplication | Tandem | X:32299171-32323344 | Pathogenic |
| DMD-8 | F | Rule out cystic fibrosis | GeneTitan genotyping microarray, exons 31–40 duplication | Tandem | X:32342897-32400997 | Pathogenic |
| DMD-9 | F | Follow-up positive carrier screening | MLPA, exons 49–50 duplication | Interspersed | X:28281185-28416485<br>X:31774879-31863353 | Likely benign |
| DMD-10 | F | Short stature | CytoScan HD microarray, arr[hg38] Xp22.11(22,134,158-22,420,550)x3,Xp21.1(31,545,342-31,835,875)x3<sup>2</sup> | Interspersed | X:22131339-22426104<br>X:31548099-31831899 | Likely benign |
| DMD-11 | F | Developmental delay | CytoScan HD microarray, arr[hg38] Xp22.33(359,510-387,877)x3,Xp21.2(31,180,559-31,262,867)x3<sup>3</sup> | Interspersed | X:341433-388232<br>X:31185540-31263036 | Likely pathogenic |
| DMD-12 | F | Follow-up positive carrier screening | MLPA, exons 52–57 duplication | Interspersed | X:31483335-31743460<br>X:33433937-33542260 | Uncertain significance |
| DMD-13 | F | Congenital heart disease | CytoScan HD microarray, arr[hg38] Xp21.1(31,853,294-32,086,953)x3,Xp21.1(32,667,715-32,898,823)x3 | Interspersed | X:31851822-32087574<br>X:32667138-32667164<br>X:32667169-32894518 | Likely pathogenic |
| DMD-14 | M | Developmental delay | MLPA, exons 5–7 duplication | Interspersed | X:32720029-32831305<br>X:120038411-120147853 | Likely benign |
| DMD-15 | Prenatal | Parent with positive carrier screening | MLPA, exons 3–4 duplication | Interspersed | X:32839747-32890599<br>Uncertain 9qh breakpoint<sup>4</sup> | Likely benign |

<sup>1</sup> In all cases, the exon contents of the intragenic *DMD* duplications revealed by long-read sequencing were consistent with that detected by the clinical assay. <sup>2</sup> This case also had another duplication, X:7,049,133-7,323,414 detected by microarray; however, it was not related to the *DMD* structural variant identified by this study. <sup>3</sup> The X:359,510-387,877 duplication was detected by microarray but not clinically reported, as it did not meet reporting criteria. <sup>4</sup> Due to the polymorphic nature of the 9qh region among individuals, the exact breakpoints within 9qh could not be determined. Abbreviations: d: days; m: months; y: years; MLPA: multiplex ligation-dependent probe amplification.

In all four cases with a clinical indication of dystrophinopathy (DMD-1 to DMD-4), the duplications were confirmed to be in tandem (Figures 1A–D). This finding aligned with expectations, as pathogenic intragenic *DMD* duplications are known to occur in tandem.^6^

**Figure 1.**
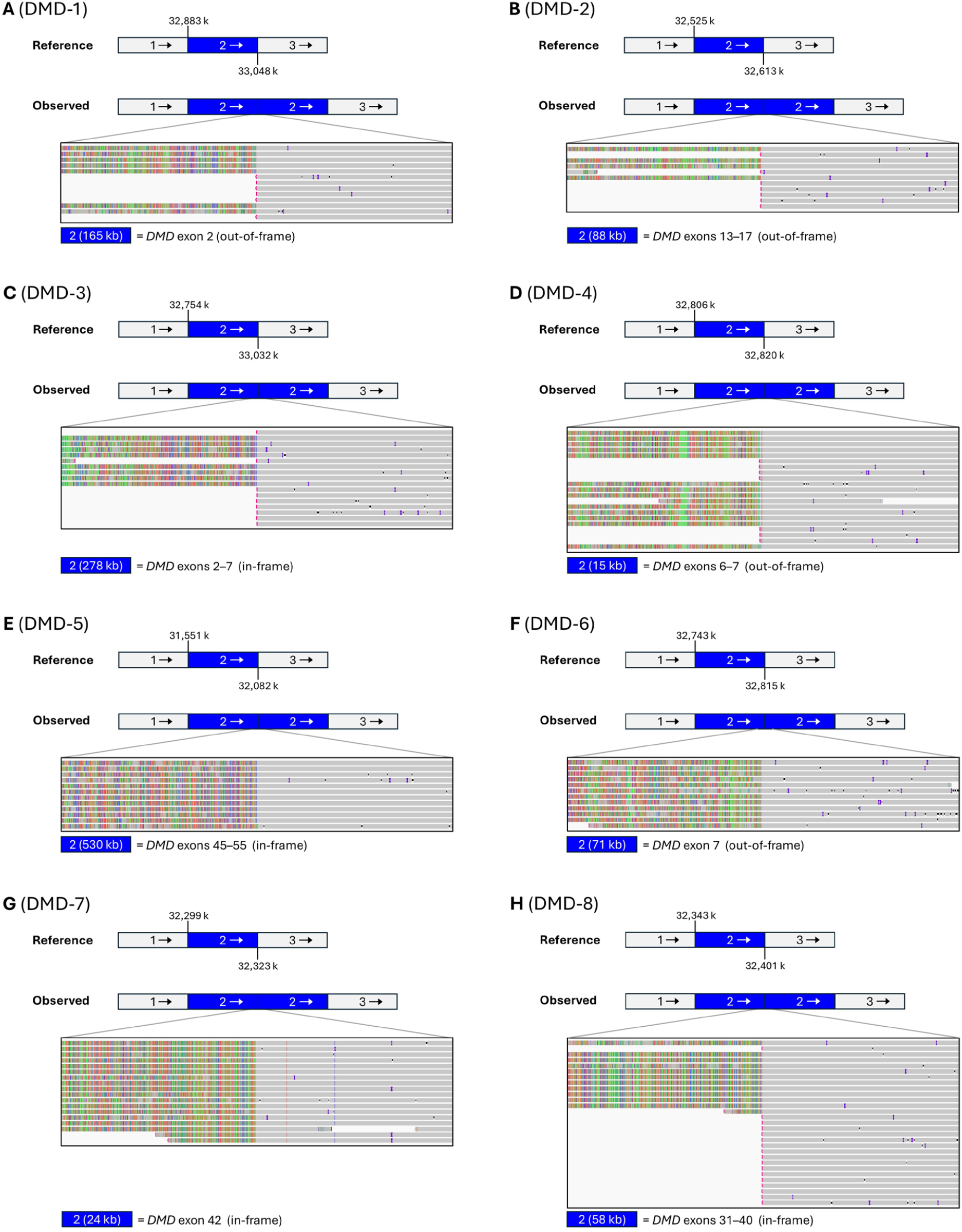
Eight cases of tandem intragenic *DMD* duplications. The blue-colored regions denote duplications. The arrows indicate orientation of the genomic segments. (A–D) Cases with a personal or family history of dystrophinopathy. (E–H) Cases with incidentally detected duplications.

Strikingly, only four of the 11 cases with incidentally detected intragenic *DMD* duplications were discovered to be in tandem (DMD-5 to DMD-8; Figures 1E–H). For three of the four cases, the duplicated exon contents had been previously reported in dystrophinopathy patients.^11,12^ The fourth case (DMD-8) harbored a duplication of exons 31–40, which had not been described before. Given that these duplications were confirmed to be in tandem, they are consistent with carrier status for dystrophinopathy. The remaining seven (64%) of the 11 cases (DMD-9 to DMD-15) were found to harbor interspersed duplications. The duplicated exon content varied across the cases (Table 1), which suggests that they arose from distinct mutagenic events.

Interestingly, four cases (DMD-9 to DMD-12) with interspersed duplications had “co-duplication” of *DMD* exons with another locus (Figure 2). The co-duplicated locus varied among the four cases but was consistently on the short arm of the X chromosome, 1.7–30.8 Mb from the duplicated *DMD* exons. In these cases, split reads and read depth supported the presence of a “co-duplication cassette” containing the duplicated *DMD* exons fused with the co-duplicated locus, in the same orientation. However, split reads alone could not resolve whether this cassette was inserted within the *DMD* gene or into the region where the co-duplicated locus originated. These two possibilities may be differentiated using long-range phasing across the cassette (illustrated in Figure 2A).

**Figure 2.**
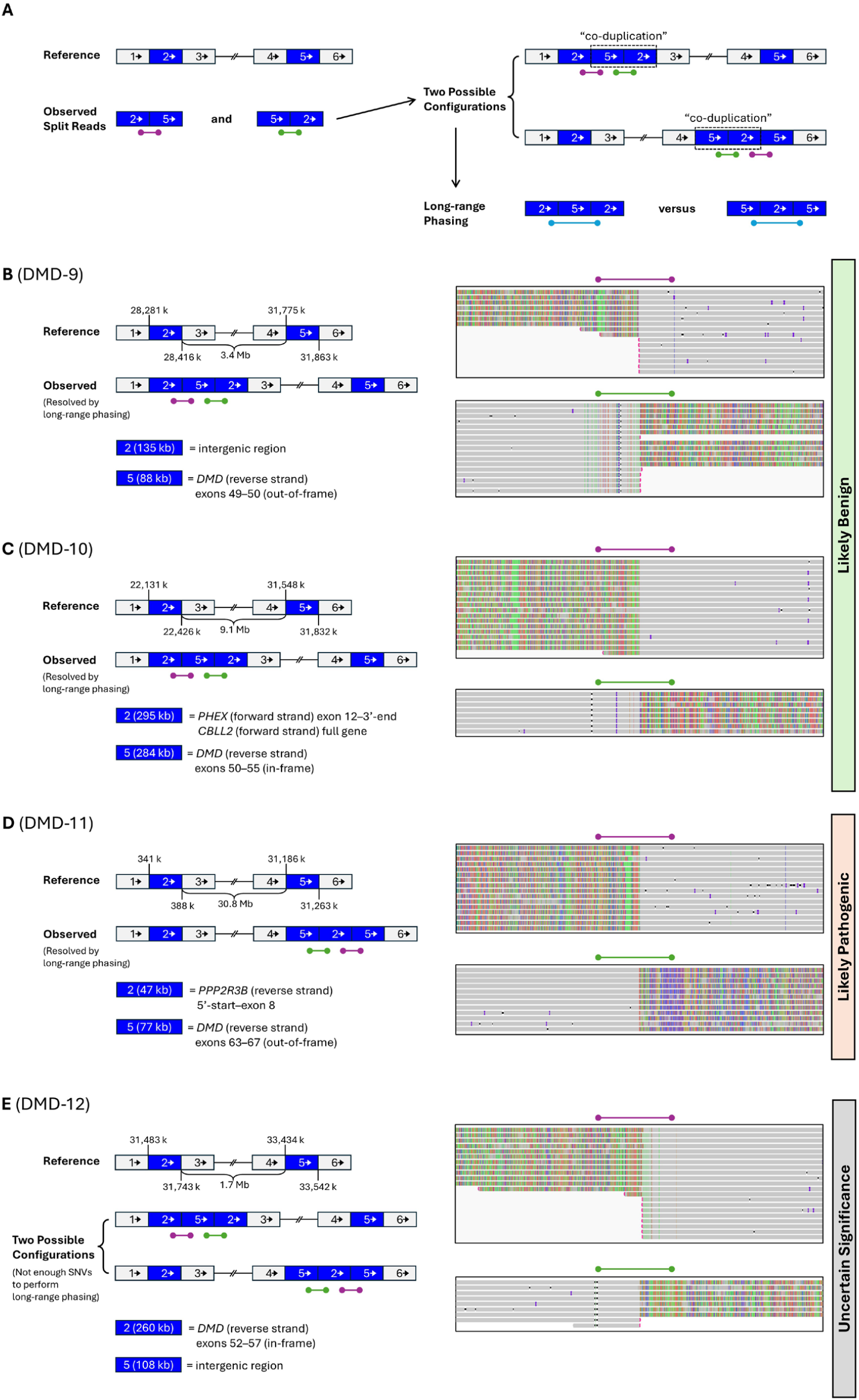
Four cases of interspersed intragenic *DMD* duplications characterized by co-duplication events. The blue-colored regions denote duplications. The arrows indicate orientation of the genomic segments. (A) Schematic overview of the co-duplication events, with two possible structural configurations of the co-duplication cassette, supported by split reads (purple and green) and copy number profiles. (B–E) Detailed co-duplication patterns in DMD-9 to DMD-12. Each case involves the duplication of specific *DMD* exons (region 5 in panels B–D and region 2 in panel E) alongside a region from the short arm of the X chromosome, located at varying distances from *DMD* (region 2 in panels B–D and region 5 in panel E). Long-range phasing using SNVs resolved the structural configuration in DMD-9 to DMD-11 (panels B–D), while the configuration remained unresolved in DMD-12 due to insufficient SNV data (panel E).

DMD-9 involved the co-duplication of an 88-kb region containing *DMD* exons 49–50 along with a 135-kb region originating 3.4 Mb telomeric to the *DMD* duplication harboring no RefSeq genes. DMD-10 involved the co-duplication of a 284-kb region containing *DMD* exons 50–55 along with a 295-kb region originating 9.1 Mb telomeric to the *DMD* duplication containing part of *PHEX* (exon 12 to the 3’-end) and the full *CBLL2* gene, both of which are located on the opposite strand relative to *DMD. PHEX* is associated with X-linked dominant hypophosphatemic rickets, while *CBLL2* is not currently associated with any diseases. In both cases, long-range phasing using single-nucleotide variants (SNVs) clarified that the co-duplication cassettes inserted into the regions where the co-duplicated loci originated (i.e., an intergenic region in DMD-9, and adjacent to *PHEX* and *CBLL2* in DMD-10; Figures 2B, 2C, S1, S2), and did not disrupt the reading frame of any genes. Therefore, we concluded that the intragenic *DMD* duplications in DMD-9 and DMD-10 were likely benign.

DMD-11 involved the co-duplication of a 77-kb region containing *DMD* exons 63–67 along with a 47-kb region originated 30.8 Mb telomeric to the *DMD* duplication and includes part of *PPP2R3B* (5’-start to exon 8). *PPP2R3B* is not currently associated with any diseases. In contrast to DMD-9 and DMD-10, long-range phasing revealed that the co-duplication cassette had inserted within *DMD* (Figures 2D and S3). Thus, the duplicated *DMD* and *PPP2R3B* (both on the reverse strand) exons may affect the reading frame of *DMD*, likely indicative of true carrier status for dystrophinopathy, although the exact impact on the *DMD* reading frame is uncertain due to the inclusion of exons from *PPP2R3B*.

DMD-12 was found to have co-duplication of a 260-kb region containing *DMD* exons 52–57 along with a 108-kb region originating 1.7 Mb centromeric to the *DMD* duplication containing no RefSeq genes. Due to the paucity of SNVs, long-range phasing was not possible for this sample. Therefore, the insertion location of the co-duplication cassette could not be determined, and this duplication remains classified as a variant of uncertain significance (VUS; Figure 2E).

In DMD-13, previous CMA testing detected duplications of *DMD* exons 3–9 and exons 45–48 (region 6 and 2, respectively, in Figure 3A). Long-read sequencing additionally identified a 27-bp intronic duplication (region 4 in Figure 3A). Split-read analysis revealed that the duplication cassette included regions 2 and 4 in an inverted orientation, fused with region 6 in the direct orientation, and that the cassette had inserted within the *DMD* gene (Figures 3A and S4). The net effect of this variant is the duplication of exons 3–9 (region 6) in the *DMD* reading frame, which had been previously reported in DMD patients.^13^ Therefore, this variant was classified as likely pathogenic, consistent with carrier status for a *DMD*-related disorder.

**Figure 3.**
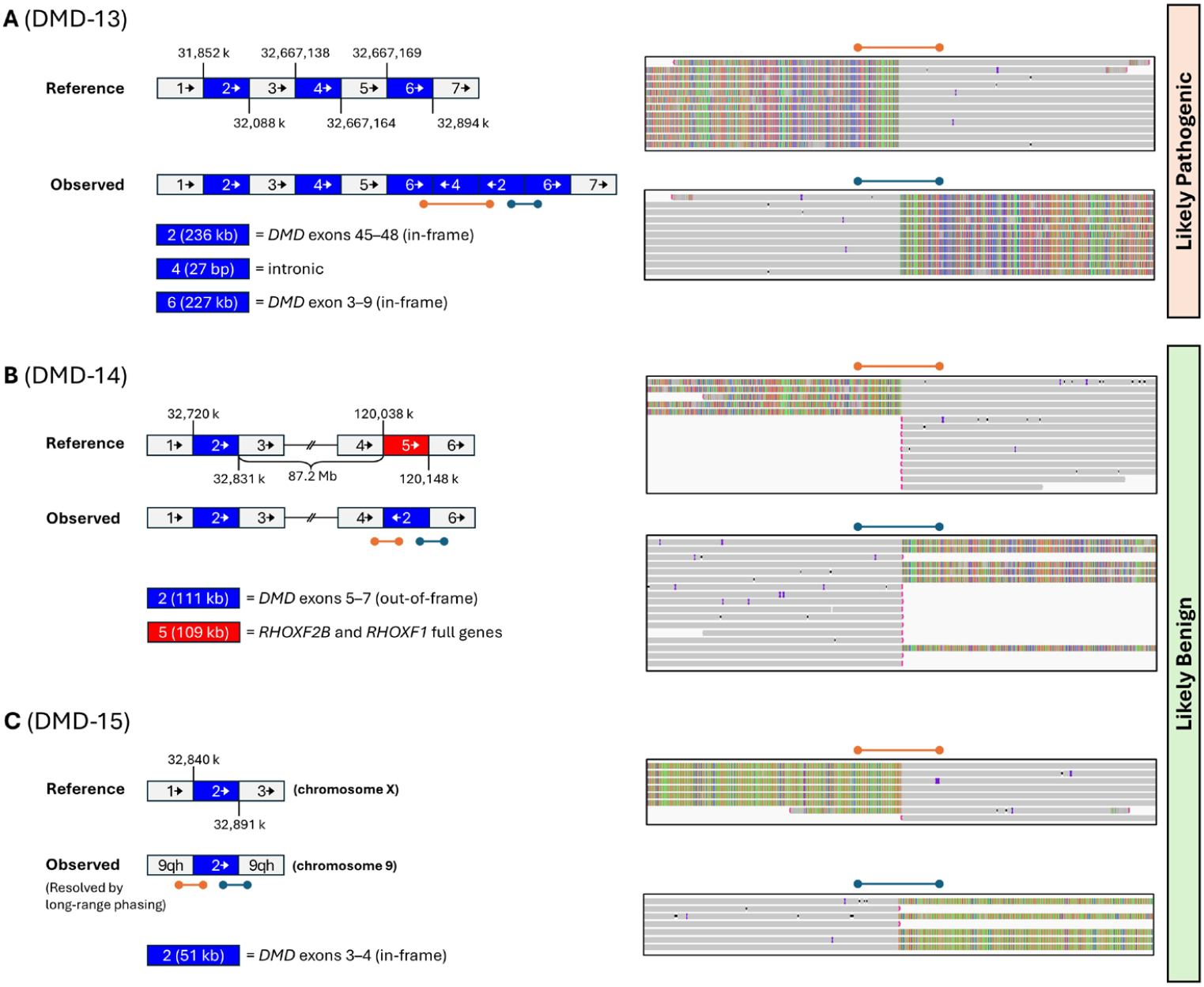
Three additional cases of interspersed intragenic *DMD* duplications. The blue- and red-colored regions denote duplications and deletions, respectively. The arrows indicate orientation of the genomic segments. (A) A complex variant involving the duplication of three distinct segments within *DMD*. (B) Duplicated *DMD* exons 5–7 inserted 87.2 Mb away on the long arm of the X chromosome, in an inverted orientation, at the site of a 109-kb deletion. (C) Duplicated *DMD* exons 3–4 inserted into the repetitive 9q12 (9qh) region.

In DMD-14, long-read sequencing revealed that the duplication of *DMD* exons 5–7 had inserted about 87.2 Mb away from their original location in an inverted orientation. At the site of the insertion, there was also a 109-kb deletion that included *RHOXF2B* and *RHOXF1*, two genes with no known disease association (Figure 3B). Because this variant did not affect the *DMD* reading frame, it was classified as likely benign.

In DMD-15, long-read sequencing revealed that the duplicated *DMD* segment containing exons 3–4 was flanked on both sides by repetitive sequences from 9q12 (i.e., the 9qh region; Figure 3C). Based on long-range phasing, we concluded that the duplicated *DMD* exons 3–4 had inserted into 9q12. Therefore, this variant was also classified as likely benign.

We also evaluated the performance of DMDuper in automatically determining the structural configuration of the duplications (i.e., tandem vs. interspersed). DMDuper achieved 100% concordance with the results from manual review (examples in Figure S5).

## Discussion

### Clinical Significance of Our Findings

Intragenic duplications of *DMD* have been extensively studied, but most of these studies have focused on patients affected by dystrophinopathy. Few, if any, systematic studies have specifically addressed patients with “incidentally detected” (i.e., asymptomatic individuals with no known family history) intragenic *DMD* duplications, despite the increasing relevance of this issue in pre-conception and prenatal genetic counseling. Our study is the largest to date to examine this group of patients. The study design aimed to minimize ascertainment bias in the group of incidentally detected duplications by selecting samples based solely on specimen availability (5 µg of high-molecular-weight DNA), without any additional criteria.

Previous reports have noted that most (~80%) of duplications throughout the genome are tandem and in direct orientation^14^. Strikingly, we found that 64 percent (7/11) of cases in our incidental detection cohort had interspersed duplications, of which four were subsequently classified as likely benign, two were classified as likely pathogenic, and one remained VUS due to the inability to fully resolve its structural configuration. Notably, had the current guidelines on the tandem presumption been followed, all of these variants would have been classified as pathogenic or likely pathogenic. Thus, our findings underscore the necessity of a more cautious and nuanced approach when interpreting intragenic *DMD* duplications detected without a clinical indication of dystrophinopathy.

Previous case reports have detected interspersed intragenic *DMD* duplications and likewise note the complexities of interpreting such variants in this gene. Bai et al.^15^ reported a male individual with *DMD* exons 56–61 duplication who was asymptomatic. PacBio long-read sequencing revealed that the duplication was interspersed, inserted over 4 Mb away from its original location. Two recent conference abstracts^16,17^ described four cases of interspersed duplications involving *DMD* exons 45–51 inserted into 17q21.1, in patients without a clinical indication of dystrophinopathy. It is important to note that both exons 45–51 and 56–61 duplications have been reported in dystrophinopathy patients (presumably in tandem), suggesting that exon content alone cannot reliably distinguish between tandem and interspersed duplications. Our study, with a larger sample size, supports the substantial prevalence of interspersed configurations among incidentally detected intragenic *DMD* duplications.

### *The Advantages of Long-read Sequencing for* DMD *Duplication Analysis*

An important methodological question is whether short-read next-generation sequencing (SR-NGS), when it covers all exons and introns of *DMD*, is sufficient to resolve the structural configuration of intragenic *DMD* duplications, or if long-read sequencing is necessary. We argue that long-read sequencing is highly advantageous over SR-NGS. Firstly, many of the interspersed duplications identified in this study have breakpoints within repetitive regions defined by RepeatMasker, which would be difficult to align and analyze accurately with short reads. This limitation of SR-NGS is best exemplified by DMD-9. This patient was initially identified as positive for an intragenic *DMD* duplication by carrier screening, and subsequently underwent a targeted SR-NGS clinical test at an external laboratory using a custom-designed *DMD* capture probeset. Despite capturing all *DMD* exons and introns (including the breakpoint regions identified by our study), SR-NGS was unable to resolve the duplication breakpoints in DMD-9, and the variant was interpreted as pathogenic by the external laboratory. This result had prompted chorionic villus sampling to determine the genotype in the patient’s male fetus—fortunately, the fetus did not carry the duplication.

Using long-read sequencing, we were able to reveal that the duplicated exons in DMD-9 were inserted into an intergenic region outside *DMD*, suggesting that the variant is in fact likely benign (Figure 2B), and the invasive prenatal testing may not have been indicated.

In addition, long-read sequencing enabled the identification of a co-duplication cassette in several cases, where *DMD* exons were duplicated alongside another locus (see Results). If the co-duplication cassette had inserted into the region where the co-duplicated locus originated (as in DMD-9 and DMD-10), it is unlikely to affect the *DMD* reading frame. However, if the cassette had inserted into the *DMD* gene itself (as in DMD-11), it may disrupt the *DMD* reading frame and be likely pathogenic. Long-range phasing across the co-duplication cassette is necessary to distinguish between these two scenarios, which is possible with long-read sequencing but not with SR-NGS.

### Towards a More Cautious Variant Interpretation Approach

This study has important implications for variant interpretation of intragenic *DMD* duplications detected in patients with no known personal nor family history of dystrophinopathy, including those undergoing carrier screening. For variants detected by an assay that cannot resolve structural configuration (i.e., “dosage-only”), such as MLPA, CMA, and most NGS assays, the laboratory should reconsider the presumption that intragenic duplications are in tandem. This presumption is inaccurate in 64% of the incidentally detected intragenic *DMD* duplications we examined. Without further information, we believe it is most prudent to classify incidentally detected intragenic *DMD* duplications as VUS until proven otherwise.

We also recommend considering long-read sequencing (or less optimally, a SR-NGS assay that sequences all exons and introns in *DMD*), when available, to be used to clarify the structural configuration of the duplication. Cytogenetic assays, e.g., fluorescence *in situ* hybridization (FISH), may assist in clarifying the location of the duplication.^16^ If the duplication was found to be in tandem, PVS1 (out-of-frame)^7^, or PVS1 with modified strength or PM4 (in-frame)^8^ may be applied. On the other hand, if the duplication was found to be interspersed, it is important to assess the impact on the *DMD* reading frame. If the duplication does not impact the reading frame of *DMD* or any other disease-causing genes, it may be classified as likely benign (as in DMD-9 and DMD-10). Otherwise, it may be classified as likely pathogenic (as in DMD-11). In cases where clarification of the structural configuration is impossible or inconclusive, it is crucial to carefully consider family history and perform familial variant testing to help interpret the pathogenicity of the variant. For example, detection of the variant in carefully phenotyped adult asymptomatic male relatives would support a likely benign classification. We summarized the proposed variant classification approach in Figure 4.

**Figure 4.**
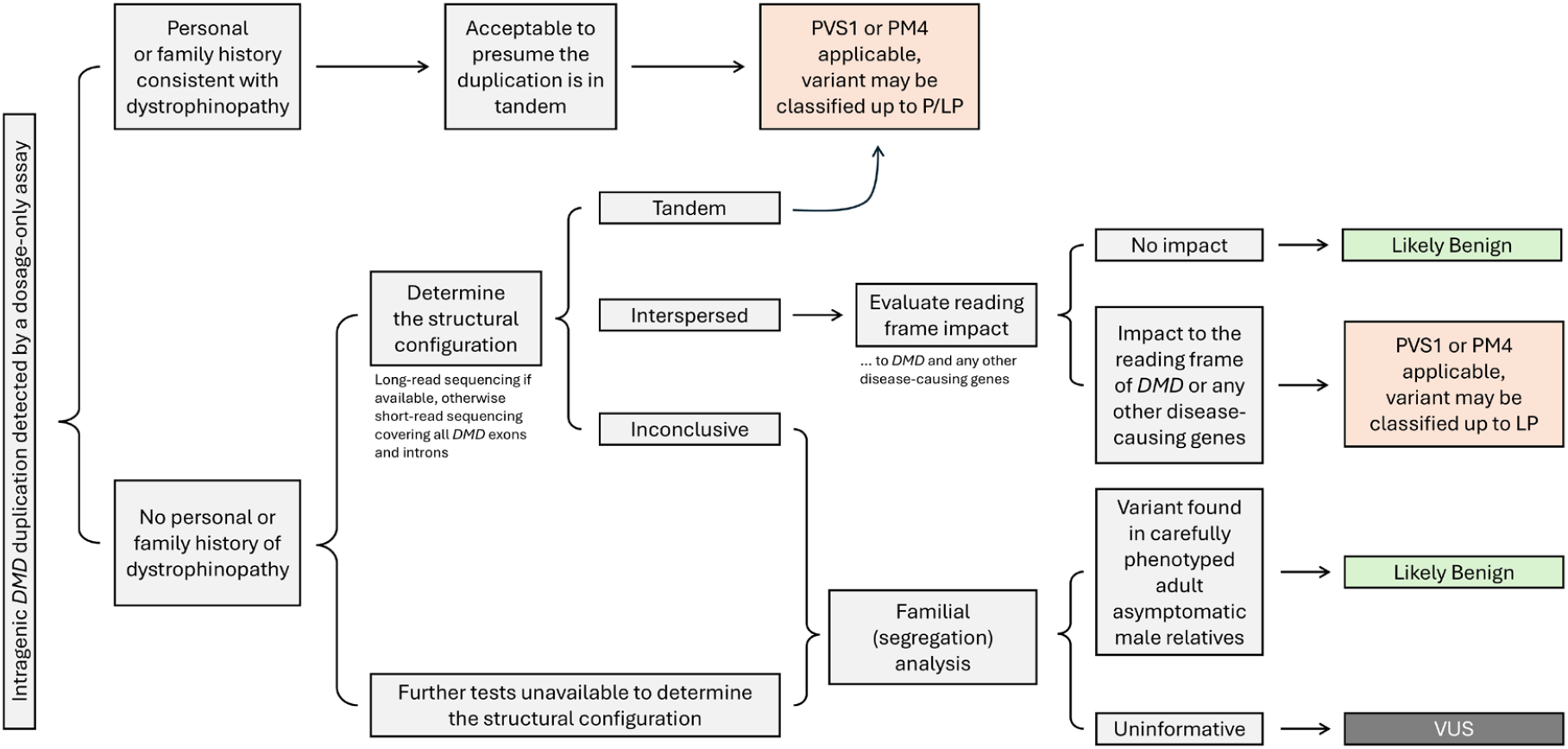
Proposed decision tree for interpretation of intragenic *DMD* duplications. This flowchart outlines the recommended steps for interpreting intragenic *DMD* duplications, emphasizing that they should not be presumed to be tandem in the absence of a clinical indication of dystrophinopathy. If additional tests to clarify the structural configuration are inconclusive or unavailable, and familial analysis is uninformative, these variants should be classified as variants of uncertain significance (VUS).

### Limitations and Future Directions

This study represents the largest investigation of the structural configurations of incidentally detected *DMD* duplications to date. However, the sample size of 15 cases is still relatively modest, and larger studies are needed to further confirm these findings. Despite the sample size, the detection of a substantial number of interspersed intragenic *DMD* duplications supports the hypothesis that these variants are not rare. We emphasize that this issue should be brought to the attention of clinical laboratory geneticists, maternalfetal medicine specialists, and prenatal genetic counselors.

## Ethics Declaration

This study was conducted as a Quality Improvement initiative at the Department of Laboratory Medicine and Pathology, Mayo Clinic, to improve interpretation of *DMD* genetic testing results. The Mayo Clinic Institutional Review Board (IRB) determined that no IRB approval was required for this project.

## Data Availability

The data generated by this study is available from the corresponding authors upon request.

## Acknowledgements

The authors thank the staff in the Department of Laboratory Medicine and Pathology, Mayo Clinic, for preparing samples and generating data used in this study. The authors also thank Dr. Sissel Juul and Dr. Nabihah Sachedina from ONT for providing administrative and technical support.

## References

1. Duan D, Goemans N, Takeda S, Mercuri E, Aartsma-Rus A. Duchenne muscular dystrophy. Nat Rev Dis Primers. 2021;7(1):13.

2. Romitti PA, Zhu Y, Puzhankara S, et al. Prevalence of Duchenne and Becker muscular dystrophies in the United States. Pediatrics. 2015;135(3):513–521.

3. Cohen G, Shtorch-Asor A, Ben-Shachar S, et al. Large scale population screening for Duchenne muscular dystrophy-Predictable and unpredictable challenges. Prenat Diagn. 2022;42(9):1162–1172.

4. Gregg AR, Aarabi M, Klugman S, et al. Screening for autosomal recessive and X-linked conditions during pregnancy and preconception: a practice resource of the American College of Medical Genetics and Genomics (ACMG). Genet Med. 2021;23(10):1793–1806.

5. Flanigan KM, Dunn DM, von Niederhausern A, et al. Mutational spectrum of DMD mutations in dystrophinopathy patients: application of modern diagnostic techniques to a large cohort. Hum Mutat. 2009;30(12):1657–1666.

6. Hu XY, Ray PN, Worton RG. Mechanisms of tandem duplication in the Duchenne muscular dystrophy gene include both homologous and nonhomologous intrachromosomal recombination. Embo j. 1991;10(9):2471–2477.

7. Abou Tayoun AN, Pesaran T, DiStefano MT, et al. Recommendations for interpreting the loss of function PVS1 ACMG/AMP variant criterion. Hum Mutat. 2018;39(11):1517–1524.

8. Brandt T, Sack LM, Arjona D, et al. Adapting ACMG/AMP sequence variant classification guidelines for single-gene copy number variants. Genet Med. 2020;22(2):336–344.

9. Jain C, Rhie A, Hansen NF, Koren S, Phillippy AM. Long-read mapping to repetitive reference sequences using Winnowmap2. Nat Methods. 2022;19(6):705–710.

10. Li H. Minimap2: pairwise alignment for nucleotide sequences. Bioinformatics. 2018;34(18):3094–3100.

11. White SJ, Aartsma-Rus A, Flanigan KM, et al. Duplications in the DMD gene. Hum Mutat. 2006;27(9):938–945.

12. Ashton EJ, Yau SC, Deans ZC, Abbs SJ. Simultaneous mutation scanning for gross deletions, duplications and point mutations in the DMD gene. Eur J Hum Genet. 2008;16(1):53–61.

13. Zamani G, Hosseinpour S, Ashrafi MR, et al. Characteristics of disease progression and genetic correlation in ambulatory Iranian boys with Duchenne muscular dystrophy. BMC Neurol. 2022;22(1):162.

14. Newman S, Hermetz KE, Weckselblatt B, Rudd MK. Next-generation sequencing of duplication CNVs reveals that most are tandem and some create fusion genes at breakpoints. Am J Hum Genet. 2015;96(2):208–220.

15. Bai Y, Liu J, Xu J, et al. Long-Read Sequencing Revealed Extragenic and Intragenic Duplications of Exons 56-61 in DMD in an Asymptomatic Male and a DMD Patient. Front Genet. 2022;13:878806.

16. Higginbotham E, Lau L, Thiruvahindrapuram B, et al. P740: DMD or not DMD? Clinical genome sequencing in the interpretation of complex copy number gains. Genetics in Medicine Open. 2024;2.

17. Guruju N, Jump V, Liu R, et al. P638: Genomic breakpoint analysis facilitates identification of complex rearrangements and re-classification of non-tandem duplications in the DMD gene. Genetics in Medicine Open. 2024;2:101544.

