## Supplementary Materials for "Rethinking the pathogenicity of intragenic *DMD* duplications detected by carrier screening: high prevalence of non-tandem duplications revealed by long-read sequencing"

**Table S1.** Sequencing metrics of the samples analyzed in this study.

| Case ID | Average autosomal coverage (fold) | N50 (bp) | Total reads | Mean read length (bp) | Percent of genome at 20x coverage |
|---|---|---|---|---|---|
| DMD-1 | 37.73 | 25,169 | 7,548,616 | 15,494 | 87.06 |
| DMD-2 | 23.70 | 19,854 | 6,483,347 | 11,600 | 71.86 |
| DMD-3 | 27.27 | 27,255 | 5,815,439 | 14,857 | 82.34 |
| DMD-4 | 38.80 | 26,865 | 8,772,611 | 13,993 | 88.54 |
| DMD-5 | 25.50 | 17,726 | 7,578,630 | 10,680 | 77.37 |
| DMD-6 | 35.92 | 19,996 | 13,608,950 | 8,401 | 88.13 |
| DMD-7 | 38.39 | 18,326 | 11,432,417 | 10,693 | 88.44 |
| DMD-8 | 39.54 | 12,037 | 18,854,076 | 6,649 | 88.53 |
| DMD-9 | 35.03 | 30,464 | 5,939,008 | 18,528 | 88.11 |
| DMD-10 | 27.99 | 41,810 | 4,553,161 | 19,404 | 82.72 |
| DMD-11 | 33.42 | 26,717 | 8,429,005 | 12,558 | 87.10 |
| DMD-12 | 28.44 | 16,662 | 18,384,287 | 4,913 | 84.66 |
| DMD-13 | 25.37 | 47,080 | 4,378,145 | 18,264 | 75.98 |
| DMD-14 | 39.81 | 10,003 | 32,224,658 | 3,854 | 87.77 |
| DMD-15 | 22.18 | 11,844 | 11,465,969 | 6,023 | 62.08 |

**DMD-9: *DMD* exons 49–50 duplicated and inserted into an intergenic region**

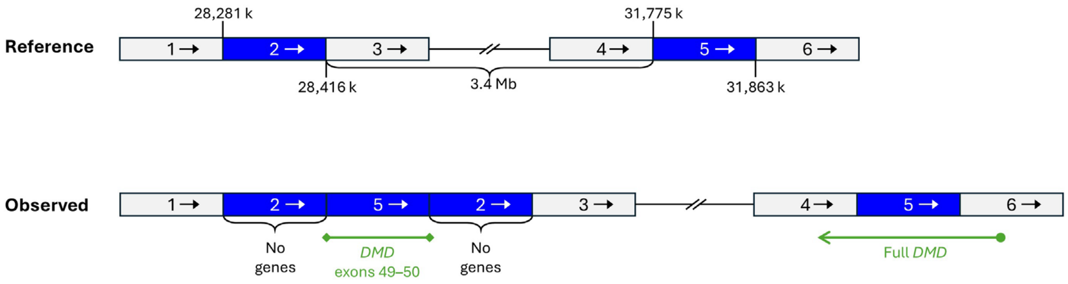


**Figure S1.** Genomic content of the intragenic *DMD* duplication in DMD-9.
The green lines in the figure represent the *DMD* gene, or parts thereof. The circle and arrow denote the 5'-start and 3'-end of a gene, respectively. The diamonds denote breakpoints within a gene. The co-duplicated region (region 2) does not contain any RefSeq genes. Since this structural variant did not disrupt the reading frame of *DMD* or any other disease-causing genes, it was interpreted as likely benign.

**DMD-10: *DMD* exons 50–55 duplicated and inserted adjacent to *PHEX* and *CBLL2***

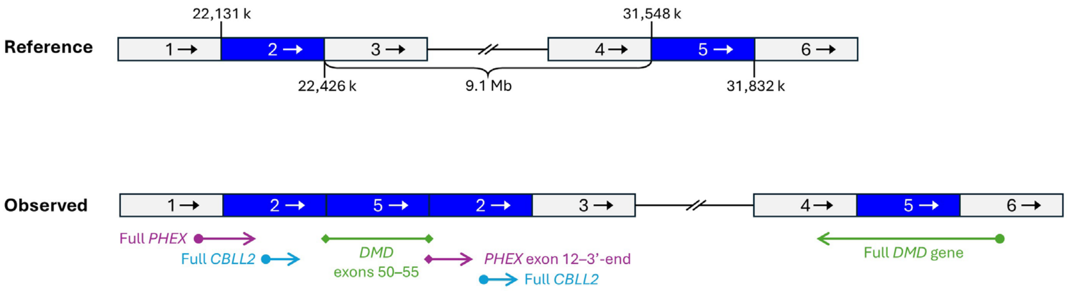


**Figure S2.** Genomic content of the intragenic *DMD* duplication in DMD-10.
The green, purple, and blue lines represent *DMD*, *PHEX*, and *CBLL2* (or parts thereof), respectively. The circle and arrow denote the 5'-start and 3'-end of a gene, respectively. The diamonds denote breakpoints within a gene. The co-duplicated region (region 2) contains exon 12 to 3'-end of *PHEX* and the full *CBLL2* gene, neither of which has any known gene-disease associations. Since this structural variant did not disrupt the reading frame of *DMD* or any other disease-causing genes, it was interpreted as likely benign.

### DMD-11: *DMD* exons 63–67 co-duplicated with *PPP2R3B* and inserted within *DMD*

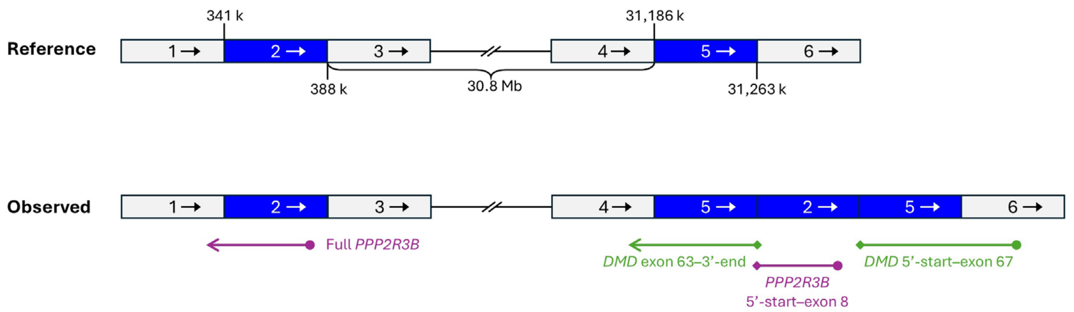


**Figure S3.** Genomic content of the intragenic *DMD* duplication in DMD-11.
The green and purple lines represent *DMD* and *PPP2R3B* (or parts thereof), respectively. The circle and arrow denote the 5'-start and 3'-end of a gene, respectively. The diamonds denote breakpoints within a gene. The co-duplicated region (region 2) contains 5'-start to exon 8 of *PPP2R3B*. This structural variant may disrupt the reading frame of *DMD*, and therefore it was interpreted as likely pathogenic.

### DMD-13: a complex structural variant involving three distinct regions within *DMD*

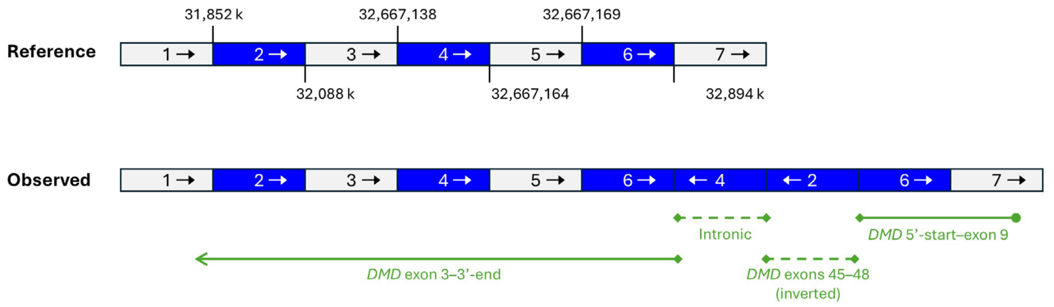


**Figure S4.** Genomic content of the intragenic *DMD* duplication in DMD-13.
The green lines in the figure represent the *DMD* gene, or parts thereof. The circle and arrow denote the 5'-start and 3'-end of a gene, respectively. The diamonds denote breakpoints within a gene. The dashed lines represent genomic segments in the inverted orientation. This complex structural variant impacts the *DMD* reading frame, and the net effect is the duplication of exons 3–9, which has been previously reported in DMD patients. For this reason, this variant was interpreted as likely pathogenic.

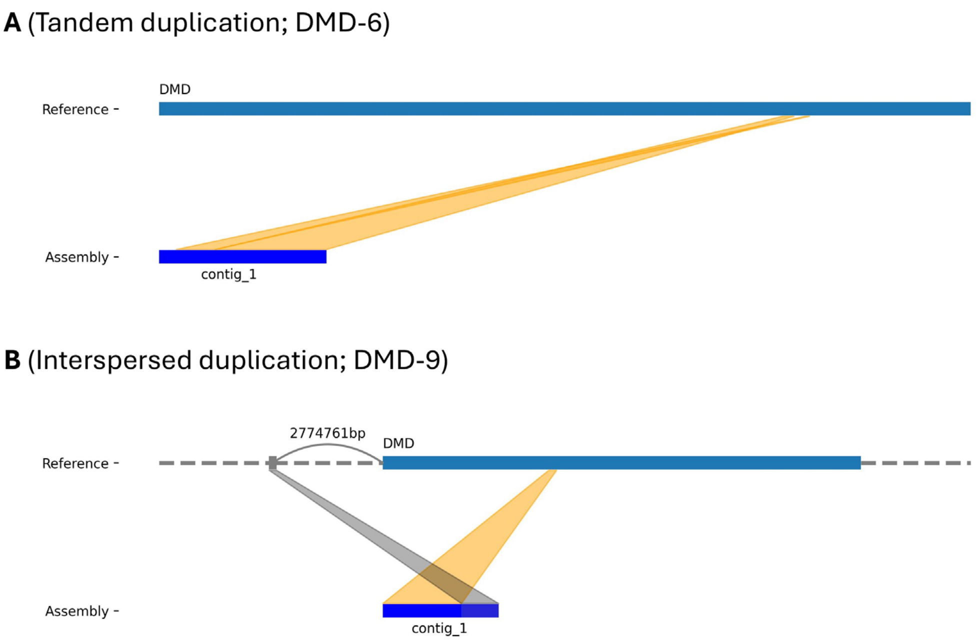


**Figure S5.** DMDuper automatically and accurately infers the structural configuration of intragenic *DMD* duplications.
(A) A case with a tandem intragenic *DMD* duplication (DMD-6). (B) A case with an interspersed intragenic *DMD* duplication (DMD-9). Note that the distance shown in panel B was calculated from the 3’-end of *DMD* and thus differs from the distance shown in Figure 2B.
